# Superior anticoagulation strategies for renal replacement therapy in critically ill patients with COVID-19: a cohort study

**DOI:** 10.1101/2020.06.26.20140699

**Authors:** Frederic Arnold, Lukas Westermann, Siegbert Rieg, Elke Neumann-Haefelin, Paul Biever, Gerd Walz, Johannes Kalbhenn, Yakup Tanriver

**Affiliations:** Department of Medicine IV: Nephrology and Primary Care, Medical Center – University of Freiburg, Faculty of Medicine, University of Freiburg, Freiburg, Germany; Institute of Medical Microbiology and Hygiene, Faculty of Medicine, University of Freiburg, Freiburg, Germany; Berta-Ottenstein-Programme for Clinician Scientists, Faculty of Medicine, University of Freiburg, Freiburg, Germany; Department of Medicine II: Division of Infectious Diseases, Medical Center – University of Freiburg, Faculty of Medicine, University of Freiburg, Freiburg, Germany; Department of Medicine III: Interdisciplinary Medical Intensive Care, Medical Center – University of Freiburg, Faculty of Medicine, University of Freiburg, Freiburg, Germany; Department of Cardiology and Angiology I, Heart Center, University of Freiburg, Germany; Department of Anesthesiology and Critical Care, Medical Center – University of Freiburg, Faculty of Medicine, University of Freiburg, Freiburg, Germany

## Abstract

**Background:** Coronavirus disease 2019 (COVID-19) patients who are admitted to intensive care units (ICU) have a high risk of requiring renal replacement therapy (RRT) due to acute kidney injury (AKI). Concomitantly, COVID-19 patients exhibit a state of hypercoagulability that can affect circuit lifespan. An optimal anticoagulation strategy is therefore needed in order to maintain circuit patency and therapeutic efficiency of RRT.

**Methods:** Retrospective single-centre cohort study on 71 critically ill COVID-19 patients at the University of Freiburg Medical Center. Included were all patients aged 18 years and older with confirmed severe acute respiratory syndrome coronavirus 2 (SARS-CoV-2) infection that were admitted to ICU between February 26 and May 21, 2020. We collected data on the COVID-19 disease course, AKI, RRT, thromboembolic events and anticoagulation. Primary outcome of the study was the effect of different anticoagulation strategies during RRT on extracorporeal circuit lifespans.

**Results:** Anticoagulation during continuous veno-venous haemodialysis (CVVHD) was performed with unfractionated heparin (UFH) or citrate. Mean treatment time in the UFH group was 21.3h (SEM: ±5.6h). Mean treatment time in the citrate group was 45.6h (SEM: ±2.7h). Citrate anticoagulation prolonged treatment duration significantly by 24.4h (*p=*0.0014). Anticoagulation during sustained low-efficiency daily dialysis (SLEDD) was performed with UFH, argatroban or low molecular weight heparin (LMWH). Mean dialysis time with UFH was 8.1h (SEM: ±1.3h), argatroban 8.0h (SEM: ±0.9h) and LMWH 11.8h (SEM: ±0.5h). Compared to UFH and argatroban, LMWH significantly prolonged treatment times by 3.7h (*p=*0.0082) and 3.8h (*p=*0.0024), respectively.

**Conclusions:** UFH fails to prevent early clotting events in dialysis circuits. For patients, who do not require an effective systemic anticoagulation, regional citrate dialysis is the most effective strategy in our cohort. For patients, who require an effective systemic anticoagulation treatment, the usage of LMWH results in the longest circuit life spans.

**Funding:** Berta-Ottenstein-Programme for Clinician Scientists, Faculty of Medicine, University of Freiburg, Germany. Else Kröner-Fresenius-Stiftung, Bad Homburg, Germany. Deutsche Forschungsgemeinschaft, Bonn, Germany.

## Introduction

The pandemic spread of SARS-CoV-2 is challenging health care systems around the world as it carries significant morbidity and mortality.^1^ Despite the predominance of respiratory symptoms, evidence emerges that COVID-19 is a multiorgan disease. SARS-CoV-2-virus copies can be detected in the kidneys, liver and brain on autopsy.^2^ AKI was registered in up to 37% of patients hospitalised for COVID-19.^3^ Endothelial and renal tropism of SARS-CoV-2 might contribute to this high incidence of AKI. Additionally, COVID-19 patients on ICU often develop a hyperinflammatory response with high levels of C-reactive protein (CRP) and excessive production of inflammatory cytokines, including Interleukin (IL)-1, IL-6, IL-8, IL-10, TNF, GM-CSF and IFNγ.^4^ This detrimental host response leads to sepsis and eventually septic shock, which will further aggravate renal function. Hence, current reports are indicating that direct and indirect effects of SARS-CoV-2 infection can lead to AKI in COVID-19 disease.

The high incidence of AKI in critically ill COVID-19 patients has led to regional shortages of dialysis supply, both in equipment and trained staff. In addition to an unusual high demand for haemodialysis, RRT has proven especially difficult due to hypercoagulability that results in intra- and extracorporeal clotting events in SARS-CoV-2 associated disease.^5^ Clotting in the extracorporeal circulation and the dialysis filter frequently requires early termination of the treatment. This reduces gross quality of the dialysis, can lead to metabolic imbalances and fluid overload, increases blood loss due to more frequent changes of the system and is likely to promote other complications for the patient. Hence, intensivists and nephrologists have acknowledged the urgent need for specific anticoagulation regimens in patients with COVID-19 requiring RRT.^6,7^

As a standard of care, critically ill patients with AKI, who require RRT, are treated either with CVVHD or SLEDD. Systemic or local anticoagulation prevents tubing and dialyzers from clotting and is therefore necessary for effective delivery of RRT. Anticoagulation during dialysis is usually maintained with UFH, which is the most commonly used anticoagulant worldwide for RRT.^8^ Alternative methods of regional and systemic anticoagulation, including citrate, LMWH (e.g. enoxaparin), heparinoids (e.g. danaparoid), thrombin antagonists (hirudin and argatroban) or platelet inhibiting agents (prostacyclin and nafamostat) have been used successfully in the past.^9^ However, their suitability in COVID-19-affected patients who require RRT have not been investigated so far.

Here, we report our experience with the use of regional citrate, LMWH and argatroban after we encountered repetitive clotting events in extra-corporal circuits of COVID-19 patients on ICU, who were previously treated with UFH. The primary goal of this study was therefore to identify the most efficient anticoagulation strategies for RRT in a representative cohort of critically ill COVID-19 patients.

## Methods

In this single-centre retrospective cohort study, we report data from the University of Freiburg Medical Center, Freiburg, which serves as a tertiary health care facility in the southwestern part of Germany. Patients eligible for inclusion were adults (aged 18 years or older) who were treated at the Medical Center with a laboratory-confirmed SARS-CoV-2 infection between February 26 and May 21, 2020.

Demographic data, past medical history, clinical findings, laboratory values, treatment details and outcome data of patients were extracted from electronic patient records by the investigators of the study (FA and LW). Follow-up data collection was continued until June 5, 2020.

For comparison, demographic, diagnostic data and outcomes of all patients (aged 18 years or older) diagnosed with influenza virus infection and admitted to an intensive care unit at the University of Freiburg Medical Center between January 1, 2015 and May 21, 2020 were extracted from electronic patient records by the investigators of the study (FA and LW). All data were reviewed and verified by two physicians (FA and LW). Any uncertain records were not included in the final data analysis.

This study is in conformity with the ethical principles for medical research involving human subjects as laid down in the Helsinki Declaration (1964) and its amendments. Analysis and publication of the data was approved by the local ethics committee (155/20 to SR and 1016/20 to JK, Chairman Prof. R. Korinthenberg).

### Laboratory procedures

Laboratory confirmation of SARS-CoV-2 infection was performed with real-time RT-PCR methods from throat-swab samples at the Institute of Virology at University of Freiburg Medical Center according to general guidelines. Concentrations of creatinine, CRP, procalcitonin (PCT), IL-6, ferritin, D-dimer, antithrombin (AT) and fibrinogen were assessed in serum samples and detected during hospitalization. Activated partial thromboplastin time (aPTT) and anti-factor Xa activity were measured for monitoring anticoagulatory therapy. Frequency of examinations was determined by the treating physician. All analyses were performed by the Institute for Clinical Chemistry and Laboratory Medicine at the University of Freiburg Medical Center. Acute kidney injury and chronic kidney disease were diagnosed according to the respective KDIGO clinical practice guidelines.^10,11^

### Haemodialysis treatments and anticoagulation

All CVVHD treatments were performed with the multiFiltrate® or the multiFiltratePRO® system (both Fresenius Medical Care GmbH, Bad Homburg, Germany) using suitable dialysate solutions, CRRT dialyzers and tubing kits. Regional citrate anticoagulation was maintained with the integrated multiFiltrate Ci-Ca® module. SLEDD treatments were performed with the GENIUS®90 system (Fresenius Medical Care GmbH, Bad Homburg, Germany) using suitable tubing kits and dialyzers for 10–12 hours. Vascular access was provided through central venous double-lumen haemodialysis catheters. Dialysate solutions were individually prepared at site. Dialysis parameters such as blood flow, dialysate flow and ultrafiltration rate were set according to system based standards and adjusted at the discretion of the treating physician.

COVID-19 patients were treated on four different ICUs at the University of Freiburg Medical Center, providing a cumulative ICU capacity of 130 beds. Every ICU is operated by an independent department (medicine, anaesthesiology, cardiovascular surgery and general surgery) and patients were randomly assigned to the different ICUs.

Therapeutic anticoagulation with UFH and argatroban were maintained by permanent intravenous infusion of the substances. Effective anticoagulation with UFH was anticipated with an activated partial thromboplastin time (aPTT) of 1.5–2.5 times the baseline, for argatroban with an aPTT of 1.5–3.0 times the baseline. Therapeutic anticoagulation with LMWH was maintained by subcutaneous injection of enoxaparin in a bodyweight adapted individual dose twice a day. Sufficient anticoagulation was supposed with an anti-factor-Xa activity of 0.5–1.0 IU/ml (measured 4 hours after injection). Before each SLEDD treatment patients received an additional intravenous bolus of 1000 I.U. enoxaparin to account for clearance during SLEDD. This protocol has been published recently.^12^

### Statistical analysis

Continuous and categorical variables were presented as median (IQR) and n (%), respectively. Treatment times were calculated as mean duration per treatment (SEM or as specified in the figure legend). Either two-sided student’s *t*-test or analysis of variance (ANOVA) with Tukey’s multiple comparisons test were applied to compare treatment times and calculate p-values. A two-sided α of less than 0.05 was considered statistically significant. All statistical analyses were performed using Prism (Version 8.0.2), GraphPad Software, San Diego, California.

## Results

Between February 26 and May 21, 2020 a total of 203 patients with COVID-19 were admitted to the University Medical Center Freiburg, of which 48 patients (24%) died. During this regional outbreak, which followed the overall pandemic development in Germany, up to 96 patients with COVID-19 had to be treated simultaneously at the University Medical Center (figure 1A). Of the 203 patients 124 were male (61%). Male patients were on average younger (62 *vs* 68 yrs) and had a more severe course of disease as indicated by ICU admission (44% *vs* 22%) or necessity for RRT (15% *vs* 5%) (figure 1B). A complete overview of the demographic and clinical characteristics of patients on ICU is shown in table 1.

**Table 1.**
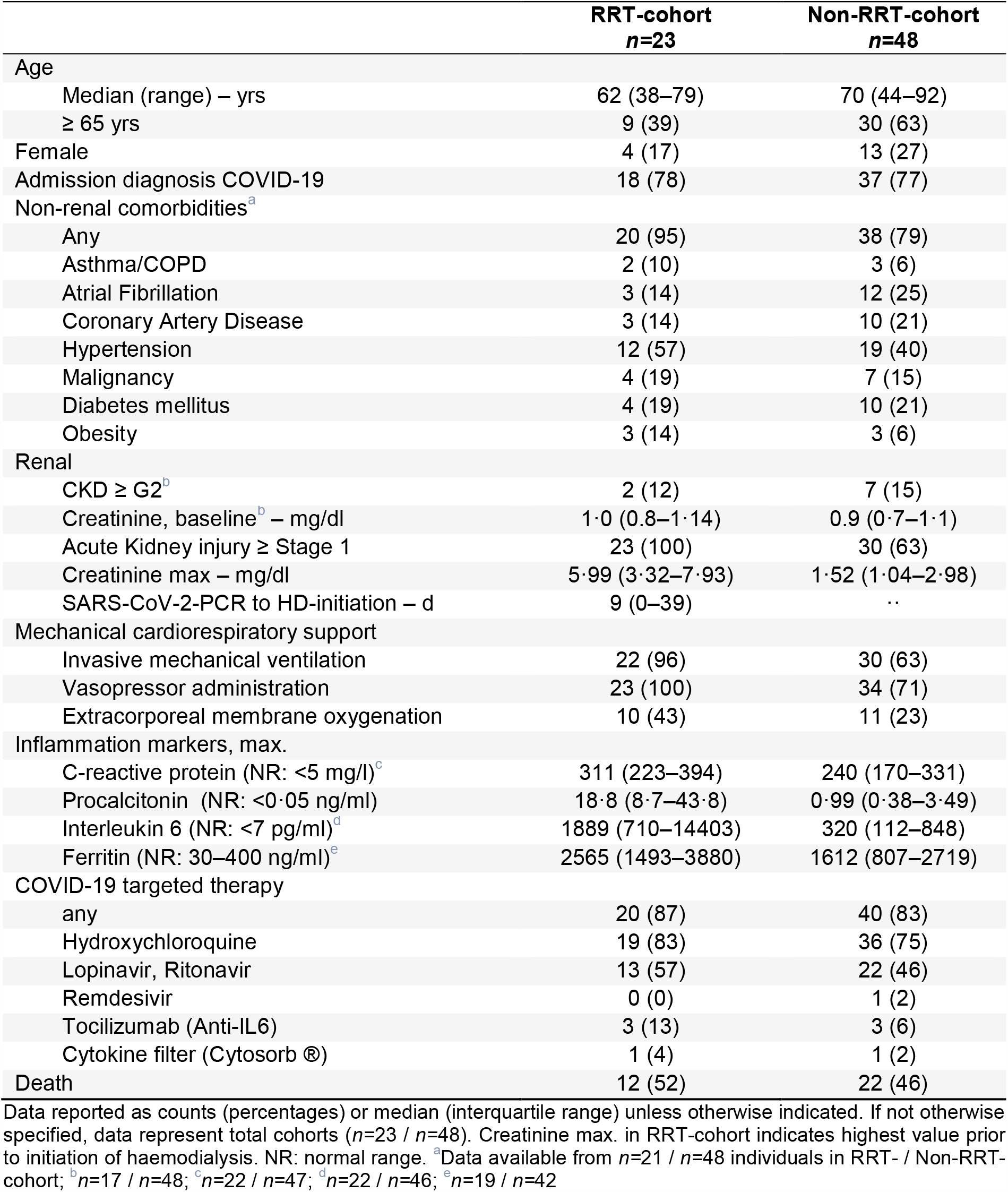
Baseline characteristics of the COVID-19 ICU-cohort

|  | RRT-cohort<br>*n*=23 | Non-RRT-cohort<br>*n*=48 |
| --- | --- | --- |
| Age |  |  |
| Median (range) – yrs | 62 (38–79) | 70 (44–92) |
| ≥ 65 yrs | 9 (39) | 30 (63) |
| Female | 4 (17) | 13 (27) |
| Admission diagnosis COVID-19 | 18 (78) | 37 (77) |
| Non-renal comorbidities<sup>a</sup> |  |  |
| Any | 20 (95) | 38 (79) |
| Asthma/COPD | 2 (10) | 3 (6) |
| Atrial Fibrillation | 3 (14) | 12 (25) |
| Coronary Artery Disease | 3 (14) | 10 (21) |
| Hypertension | 12 (57) | 19 (40) |
| Malignancy | 4 (19) | 7 (15) |
| Diabetes mellitus | 4 (19) | 10 (21) |
| Obesity | 3 (14) | 3 (6) |
| Renal |  |  |
| CKD ≥ G2<sup>b</sup> | 2 (12) | 7 (15) |
| Creatinine, baseline<sup>b</sup> – mg/dl | 1.0 (0.8–1.14) | 0.9 (0.7–1.1) |
| Acute Kidney injury ≥ Stage 1 | 23 (100) | 30 (63) |
| Creatinine max – mg/dl | 5.99 (3.32–7.93) | 1.52 (1.04–2.98) |
| SARS-CoV-2-PCR to HD-initiation – d | 9 (0–39) | .. |
| Mechanical cardiorespiratory support |  |  |
| Invasive mechanical ventilation | 22 (96) | 30 (63) |
| Vasopressor administration | 23 (100) | 34 (71) |
| Extracorporeal membrane oxygenation | 10 (43) | 11 (23) |
| Inflammation markers, max. |  |  |
| C-reactive protein (NR: <5 mg/l)<sup>c</sup> | 311 (223–394) | 240 (170–331) |
| Procalcitonin (NR: <0.05 ng/ml) | 18.8 (8.7–43.8) | 0.99 (0.38–3.49) |
| Interleukin 6 (NR: <7 pg/ml)<sup>d</sup> | 1889 (710–14403) | 320 (112–848) |
| Ferritin (NR: 30–400 ng/ml)<sup>e</sup> | 2565 (1493–3880) | 1612 (807–2719) |
| COVID-19 targeted therapy |  |  |
| any | 20 (87) | 40 (83) |
| Hydroxychloroquine | 19 (83) | 36 (75) |
| Lopinavir, Ritonavir | 13 (57) | 22 (46) |
| Remdesivir | 0 (0) | 1 (2) |
| Tocilizumab (Anti-IL6) | 3 (13) | 3 (6) |
| Cytokine filter (Cytosorb®) | 1 (4) | 1 (2) |
| Death | 12 (52) | 22 (46) |

Data reported as counts (percentages) or median (interquartile range) unless otherwise indicated. If not otherwise specified, data represent total cohorts (*n*=23 / *n*=48). Creatinine max. in RRT-cohort indicates highest value prior to initiation of haemodialysis. NR: normal range. <sup>a</sup>Data available from *n*=21 / *n*=48 individuals in RRT- / Non-RRT-cohort; <sup>b</sup>*n*=17 / *n*=48; <sup>c</sup>*n*=22 / *n*=47; <sup>d</sup>*n*=22 / *n*=46; <sup>e</sup>*n*=19 / *n*=42

**Figure 1.**
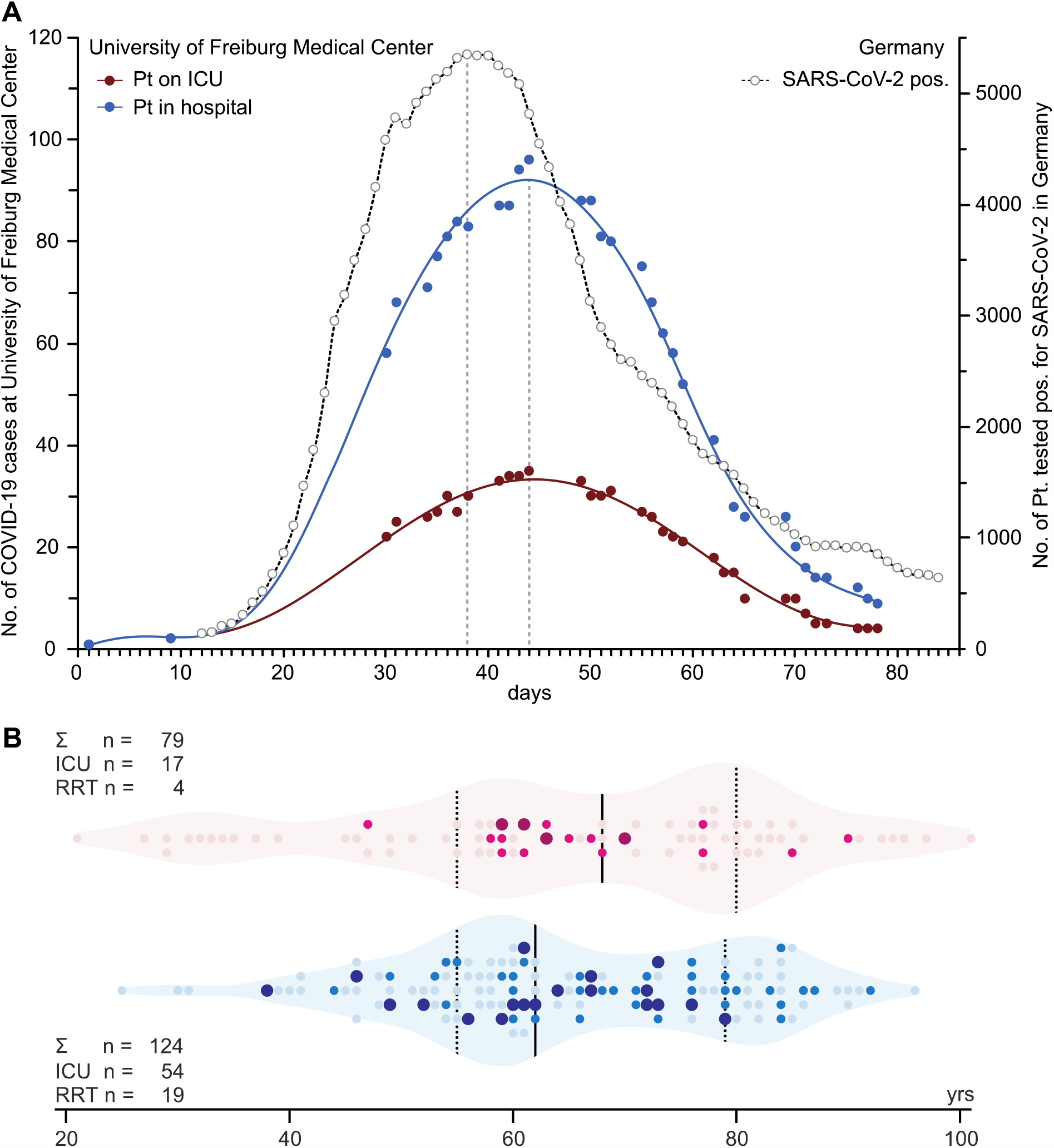
Intensive care and renal replacement therapy in COVID-19 cases at the University of Freiburg Medical Center 02/26/2020–05/21/2020 (85 d) **(A) COVID-19 cases at the University of Freiburg Medical Center 02/26/2020–05/21/2020 (85 d).** The blue graph (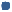) shows the total number of SARS-CoV-2 positive patients treated at the University Medical Center Freiburg at a given timepoint within a 85 day period (02/26/2020–05/21/2020) during the peak of the COVID-19 pandemic in Germany. The red graph (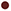) depicts the number of COVID-19 positive patients treated on an intensive care unit at University Medical Center. The dashed graph (○) shows the number of patients tested positive for SARS-CoV-2 in Germany. The temporal relation to the transmission dynamics and course of the pandemic in Germany with an approximate delay from positive testing to hospitalization of 5–7 days is demonstrated by the count of positive SARS-CoV-2 tests. (Data reported by Robert Koch Institute and local health authorities); **(B) Age distribution, ICU admission and RRT application in COVID-19-cohort**. Light dots depict all, dark dots depict individuals treated on an ICU for COVID-19. Enlarged dots mark individuals receiving RRT (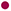 females; 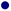 males). Median age (quartile ranges): females 68 yrs (55–80), males 62 yrs (55–79).

71 patients (35%) were treated on ICU. Of the ICU-cohort, 53 (75%) individuals developed AKI and 23 (32%) individuals were treated with RRT. Patients requiring RRT (RRT-cohort) were on average eight years younger than patients in the Non-RRT-cohort (62 *vs* 70 yrs). We also observed a male predominance in our RRT-cohort and most patients that required RRT had higher rates of pre-existing comorbidities, such as chronic obstructive pulmonary disease, hypertension and obesity. Of note, baseline renal function was not different between the two groups and most patients had no renal impairment before hospital admission. In line with their disease course, cardiorespiratory support (i.e. invasive mechanical ventilation, ECMO, vasopressors) was more often administered in the RRT-cohort. These patients showed significantly higher levels of the inflammation markers CRP, PCT and IL-6 (figure 2). Application of COVID-19 targeted therapies did not correlate with the requirement for RRT. Finally, there was a higher mortality rate in the RRT-cohort (52%) when compared to the Non-RRT-cohort (46%) (table 1 & supplement table S1). We were surprised by the high incidence of AKI, which frequently required RRT in critically ill patients diagnosed with COVID-19. The question emerged whether critically ill patients with other systemic viral infections had a similar incidence of renal complications at our hospital. Consequently, we compared the current COVID-19 ICU-cohort to a previous cohort of 200 influenza patients (including Influenza A, B) treated on ICU at the University of Freiburg Medical Center from 2015–2020 (supplement table S1). These cases represent all patients diagnosed with Influenza and admitted to the ICU (13% of total influenza cases) during this five year period.

**Figure 2.**
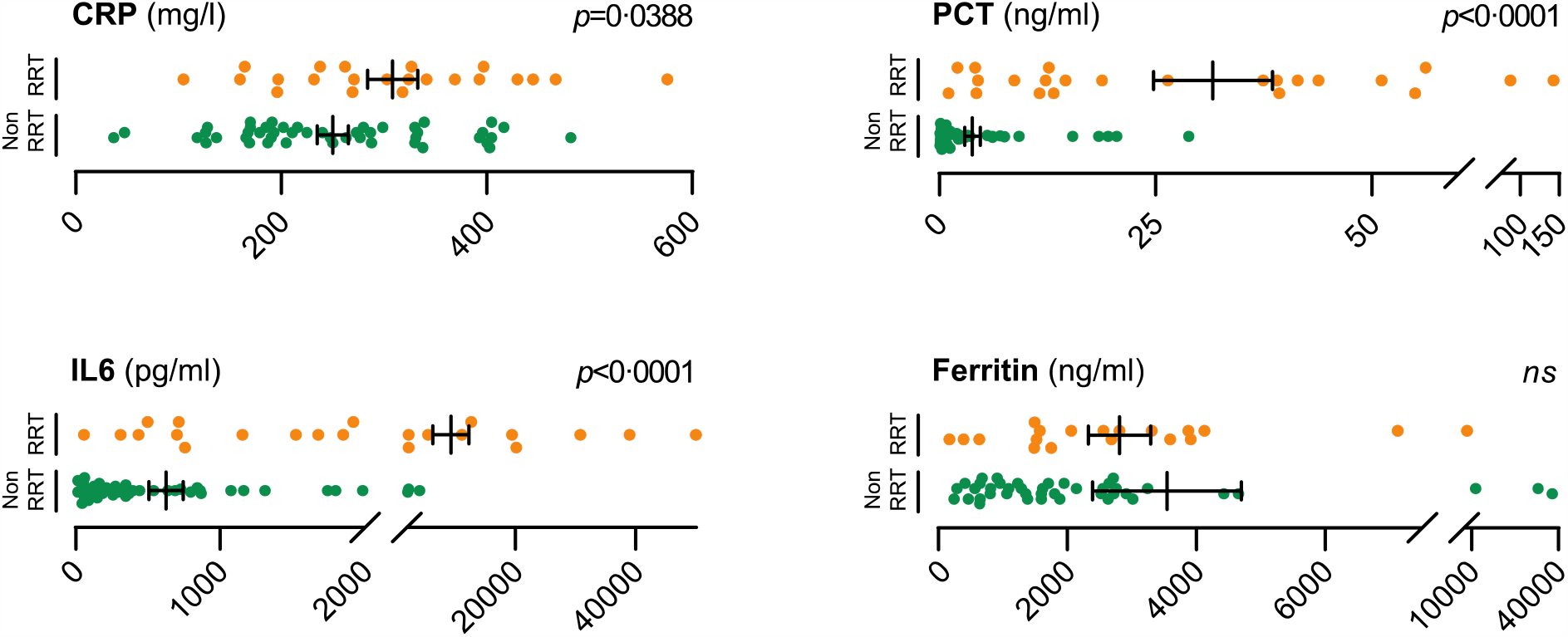
Distribution of inflammation markers in COVID-19 RRT- and Non-RRT-cohort. **Distribution of inflammation markers in COVID-19 RRT- and Non-RRT-cohort**. Dot plots show levels of selected inflammation markers CRP (**A**), PCT (**B**), IL6 (**C**) and Ferritin (**D**). Orange dots (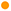) represent individual patients who underwent CVVHD or SLEDD. Green dots (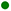) represent patients who were not treated with RRT. Bars depict mean and standard error of the mean (SEM). P-values were calculated using a two-tailed student’s *t*-test. *ns*: not significant with α=0.05. CRP available from *n=*22 / *n=*47 (RRT-/ Non-RRT-cohort); PCT *n=*23 / *n=*48; IL6 *n=*22 / *n=*46; Ferritin *n=*19 / *n=*42.

We were able to uncover that COVID-19 patients in fact have a higher demand for RRT than influenza patients (RRT-rates: 32% *vs* 19%). This might indicate a more severe course of disease in COVID-19, which was further corroborated by the higher mortality rate of patients suffering from COVID-19 compared to influenza patients (48% *vs* 24%). The observed male predominance in COVID-19 patients admitted to ICU was less pronounced in influenza patients (sex ratios: 1:3.2 *vs* 1:1.6) (supplement table S1). Overall, males required RRT more often than females during systemic viral infection, but especially when severe AKI was COVID-19 associated (supplement figure S1).

Initially, intravenous UFH was administered for systemic anticoagulation in patients who received RRT for COVID-19 related AKI. RRT was either performed as CVVHD or SLEDD. In these patients we observed frequent clotting events in the extracorporeal circulation and the dialysis filter. Our data also indicated a higher rate of systemic thromboembolic events in the RRT-cohort (35% *vs* 19%) (table 2). Supporting this finding of hypercoagulability, we also noticed a trend towards higher levels of D-dimer (8.68 mg/l *vs* 5.01 mg/l) as well as lower levels of fibrinogen (288 mg/dl *vs* 356 mg/dl) and AT (70% *vs* 76%) in the RRT-cohort (supplement figure S2).

**Table 2.** Thromboembolic events and prothrombotic markers

|  | **RRT-cohort**<br>**n=23** | **Non-RRT-cohort**<br>**n=48** |
| --- | --- | --- |
| Anticoagulation prior to hospitalization | 5 (22) | 11 (23) |
| Intracorporeal thromboembolic events | 8 (35) | 9 (19) |
| **Prothrombotic markers** |  |  |
| D-dimer max (NR: <0,5 mg/l)<sup>a</sup> | 8.68 (5.0–35.0) | 5.01 (2.30–16.72) |
| Antithrombin min (NR: 79–120%)<sup>b</sup> | 70 (61–92) | 76 (70–93) |
| Fibrinogen min (NR: 170–420 mg/dl)<sup>c</sup> | 288 (230–404) | 356 (158–564) |
| Fibrinogen max (NR: 170–420 mg/dl)<sup>d</sup> | 580 (422–673) | 559 (465–681) |

Data reported as median (interquartile range) values. For patients who underwent extracorporeal membrane oxygenation (ECMO) D-dimer max values were obtained prior to ECMO implantation. NR: normal range. <sup>a</sup>Data available n=19 / n=44 (RRT- / Non-RRT-cohort); <sup>b</sup>n=15 / n=16; <sup>c</sup>n=15 / n=15; <sup>d</sup>n=19 / n=28.

Due to frequent extracorporeal clotting events with early termination of treatments, anticoagulatory regimens were changed at the discretion of the treating physician according to the patient’s individual risk profile and departmental experience. As a result, patients receiving CVVHD were regularly switched to a regional anticoagulatory regimen with citrate. In cases where systemic anticoagulation was warranted, patients received either intravenous UFH or argatroban. Patients undergoing SLEDD were switched from UFH to alternative systemic anticoagulation with either intravenous argatroban or subcutaneously administered LMWH.

To investigate the efficiency of the different anticoagulatory regimens, we compared mean treatment times. In the context of CVVHD, 7 patients (treatments: *n=*13) were administered UFH, 18 patients (*n=*92) underwent regional anticoagulation with citrate. Mean treatment time in the citrate group was 45.6h (SEM: ±2.7h). Mean treatment time in the UFH group was 21.3h (SEM: ±5.6h). Citrate anticoagulation prolonged treatment duration significantly by 24.4h (*p=*0.0014), increasing mean treatment time more than twice. As a standard procedure time for CVVHD without change of dialysis filter or equipment we defined 48h. In the UFH group just 8% of the treatments were running for at least 48h. Compared to that, 45% of the treatments in the citrate group were running longer than 48h (table 3 & figure 3A).

**Table 3.** Counts and treatment times of continuous veno-venous haemodialysis (CVVHD) in COVID-19

|  | **No. of**<br>**treatments** | **No. of**<br>**patients** | **Total therapy**<br>**duration – h** | **Mean**<br>**duration /**<br>**treatment – h** | **Early RRT**<br>**termination –**<br>**%** |
| --- | --- | --- | --- | --- | --- |
| UFH | 13 | 7 | 274.7 | 21.1 (±20.2) | 92 |
| Citrate | 92 | 18 | 4192.6 | 45.6 (±25.6) | 55 |

Data reported as counts, mean values (standard deviation) and percentages. For the calculation of the percentage of early haemodialysis termination a minimum operating time per CVVHD treatment of at least 48h was chosen. Percentages depict portion of treatments running less than 48h.

**Figure 3.**
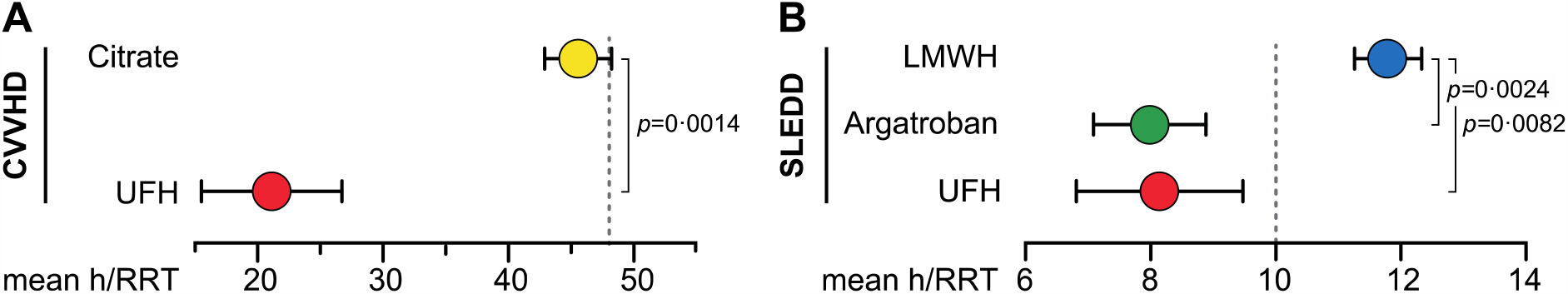
Treatment times of RRT. **Treatment times of patients receiving CVVHD (A) or SLEDD (B)**. Circles show mean duration of treatment according to anticoagulant regimen. Error bars depict SEM. P-values were calculated using a two-tailed student’s *t*-test comparing the CVVHD-subgroups and ANOVA with Tukey’s multiple comparisons test comparing the SLEDD-subgroups. Dashed lines represent minimal expected treatment times (48h for CVVHD, 10h for SLEDD).

Patients that underwent SLEDD procedures were administered systemic anticoagulation. 9 Patients (*n=*22) received UFH, 3 patients (*n=*27) argatroban and 7 patients (*n=*76) subcutaneously administered LMWH. Mean dialysis time in the UFH group was 8.1h (SEM: ±1.3h), in the argatroban group 8.0h (SEM: ±0.9h) and in the LMWH group 11.8h (SEM: ±0.5h). Use of LMWH significantly prolonged treatment times by 3.7h (*p=*0.0082) and 3.8h (*p=*0.0024) compared to the UFH group and argatroban group, respectively. Postulating a standard treatment time of 10h for SLEDD procedures, 64% of treatments in the UFH group and 67% in the argatroban group had to be terminated early, whereas just 30% of treatments in the LMWH group were conducted for less than the anticipated 10h. Hence, systemic anticoagulation with LMWH in COVID-19 patients receiving SLEDD results in a significant reduction of thromboembolic events in the extracorporeal circulation and permits sufficient dialysis in the majority of patients (table 4 & figure 3B).

**Table 4.** Counts and treatment times of sustained low-efficient daily dialysis (SLEDD) in COVID-19

|  | **No. of**<br>**treatments** | **No. of**<br>**patients** | **Total therapy**<br>**duration – h** | **Mean**<br>**duration /**<br>**treatment – h** | **Early RRT**<br>**termination –**<br>**%** |
| --- | --- | --- | --- | --- | --- |
| UFH | 22 | 9 | 179.1 | 8.1 (±6.2) | 64 |
| Argatroban | 27 | 3 | 215.6 | 8.0 (±4.7) | 67 |
| LMWH | 76 | 7 | 896.4 | 11.8 (±4.7) | 30 |

Data reported as counts, mean values (standard deviation) and percentages. For the calculation of the percentage of early haemodialysis termination a minimum operating time per SLEDD treatment of at least 10h was chosen. Percentages depict portion of treatments running less than 10h.

In conclusion, our data show that patients with COVID-19 who are admitted to ICU often develop AKI that demands RRT. Patients that require RRT have significantly elevated levels of inflammation markers and tend to develop a more severe course of disease. As part of their multiorgan disease, these patients also suffer from aggravated hypercoagulability with a high risk for intra- and extracorporeal thromboembolic events. We were able to show that regional citrate anticoagulation for CVVHD and systemic LMWH anticoagulation for SLEDD are superior anticoagulatory regimens that reduce clotting events and allow significantly longer treatment times.

## Discussion

Regional outbreaks of SARS-CoV-2 pose huge challenges to health care providers since transmission dynamics lead to rapid and simultaneous admission of large numbers of moderate to critically ill patients. Mortality of critically ill patients is high compared to other pandemic viral diseases such as influenza (48% *vs* 24% in this study).^13-15^ Besides the early predominance of respiratory symptoms COVID-19 can quickly develop into a multiple organ dysfunction syndrome, which often involves the kidneys.^2,16^ As a result critically ill patients suffering from COVID-19 more often required RRT than patients suffering from influenza (32% *vs* 19%).

This report depicts a single-centre experience with a cohort of 71 critically ill COVID-19 patients and primarily aims to give useful practical advice to healthcare professionals during an ongoing global crisis and in view of potential future outbreaks. Due to high risk for AKI, SARS-CoV-2-outbreaks confront health care providers with an unusual high demand for haemodialysis infrastructure. Hypercoagulability in critically ill patients with COVID-19 complicates RRT by significantly reducing treatment times. Premature termination due to clotting events increases blood loss, causes electrolyte, and acid-base disturbances and complicates volume management.^17-19^ Early termination of RRT increases workload and aggravates potential shortages in both staff and dialysis supplies.^20^ Evidence-based anticoagulation strategies in this subgroup of COVID-19 patients ensuring efficient RRT are therefore urgently needed. With this study we propose superior anticoagulation regimens for critically ill COVID-19 patients requiring RRT.

In our RRT-cohort, we observed unusually short treatment times for CVVHD and SLEDD in patients who were anticoagulated with UFH. First, 92% of CVVHD treatments had to be discontinued prematurely despite systemic anticoagulation with UFH. Second, administration of UFH in patients receiving SLEDD resulted in early treatment termination due to extracorporeal clotting in 64% of the cases. The anticoagulatory effect of UFH is conveyed by binding and potentiating the inhibitory actions of AT. The formation of an UFH-AT-complex inhibits mainly factor Xa and thrombin (factor IIa). Hence, reduced bioavailability of administered UFH, overwhelming activation of the coagulation cascade, as well as hereditary or acquired AT deficiency can all hamper the efficiency of UFH.^21^

Severe COVID-19 is characterized by an excessive production of inflammatory proteins, as shown here and in other studies.^4^ Several acute-phase proteins have been demonstrated to bind and inactivate UFH, thereby limiting its anticoagulatory effects.^22^ Furthermore, we noticed a significant increase in prothrombotic proteins indicating hypercoagulability, which goes in line with the reported high incidence of thromboembolic events in COVID-19 patients.^23^ High concentrations of prothrombotic factors could result in an increased fraction of functional factor Xa and thrombin, favouring cleavage of fibrinogen and leading to clotting events. Ongoing coagulation and thrombus formation reduces circulating AT, which was most noticeable in our RRT-group. Lastly, age has been shown to be a prognostic marker for UFH resistance.^24^ In the RRT-group, 39% of the patients were 65 years or older and therefore at risk for UFH resistance.

In patients with SLEDD, the LMWH enoxaparin significantly improved mean duration time in comparison to UFH and argatroban. The cause for this finding remains to be determined. However, there are factors which could contribute to superior anticoagulatory effects of LMWH. In contrast to UFH, application of LMWH does not lead to depletion of tissue factor pathway inhibitor (TFPI), which is a potent inhibitor of the extrinsic coagulation pathway that is triggered by tissue factor.^25^ Additionally, LMWH has a longer half-life, better bioavailability and lower affinity for plasma protein binding sites than UFH, which all contribute to a more reliable anticoagulation.^9^ This is further corroborated by a recent study that demonstrated a survival benefit when COVID-19 patients with a high concentration of D-dimers or sepsis-induced coagulopathy were treated with LMWH.^26^

Administration of citrate significantly improved mean treatment duration of CVVHD for COVID-19 patients. Regional anticoagulation with citrate is a suitable alternative to UFH in critically ill patients during CVVHD.^27,28^ In contrast to systemic anticoagulation, citrate complexes calcium. At a concentration of 4–6 mmol/l with an ionized calcium of <0.2 mmol/l citrate prevents activation of both coagulation cascades and platelets. These widespread effects very well explain its superior ability to prevent extracorporeal clotting events. Since citrate provides regional anticoagulation, prevention of systemic thromboembolism with citrate is not feasible. However, regional anticoagulation with citrate and additional systemic anticoagulation tailored for patients with COVID-19 will further increase patient safety compared to sole systemic anticoagulation with UFH.

An increased risk for thromboembolism due to hypercoagulability with elevation of prothrombotic markers has frequently been observed and seems to be a hallmark of severe COVID-19 disease and poor outcome.^5,23,29^ In our COVID-19 RRT-cohort, 35% of the patients developed intracorporeal thromboembolic events despite anticoagulation. It is speculated that coagulopathy in COVID-19 patients might be caused by systemic inflammation.^30^ However, recent studies also indicate a direct involvement of SARS-CoV-2 infection. Viral elements were detected in endothelial cells of multiple organs that might facilitate the induction of endotheliitis. COVID-19-endotheliitis could be a crucial factor for microvascular dysfunction.^30,31^ Significant increase of inflammation markers (e.g. CRP, PCT, IL-6) is present in patients with SARS-CoV-2 infection at admission.^32^ In our study critically ill COVID-19 patients requiring RRT showed higher inflammation markers compared to those not requiring RRT. This could further aggravate hypercoagulability and predispose to intra- and extracorporeal clotting.

Limits of our study are a short observational period and the small sample size at a single-centre. In addition, groups treated with UFH are particularly small due to frequently observed complications and early adaption of internal operating procedures recommending alternative anticoagulation strategies. However, since UFH has been the standard choice of anticoagulation during RRT for decades empirical evidence in combination with our own study clearly indicates that alternative anticoagulants might be preferable in COVID-19 associated hypercoagulability. Finally, this study includes cases from the early phase of the local outbreak. At that time no standardized laboratory workup was in place. Therefore, not all laboratory parameters were available for every patient.

In summary, our report adds to the growing body of evidence that COVID-19 can evolve into a multiorgan disease that frequently results in acute kidney injury with a high demand for RRT. Severe cases of COVID-19 are often accompanied by a state of hypercoagulability that poses a challenge to administer efficient RRT. We were able to demonstrate that mean RRT duration time can be improved significantly by using either regional anticoagulation with citrate for CVVHD or systemic anticoagulation with LMWH for SLEDD. Despite its limitations, our study data can therefore guide clinical decisions for patients requiring anticoagulation for RRT during this ongoing pandemic. Ultimately, a better understanding of COVID-19 associated hypercoagulability is required to offer tailored anticoagulation strategies to maintain extracorporeal circuit patency and reduce overall mortality from thromboembolic events.

## Data Availability

Authors had full access to the patient records, and take responsibility for the accuracy and integrity of the data.

## Contributors

FA, LW and YT conceived the study and its design, had full access to the patient records, and take responsibility for the accuracy and integrity of the data. FA and LW screened the electronic patient records, organized the data and performed statistical analysis. YT critically contributed to data analysis. FA, LW and YT drafted the manuscript. FA generated the figures. YT, JK, ENH, PB, SR, and GW were involved in clinical management of the patients, or contributed to data interpretation. All authors critically revised the drafted manuscript and approve of the submission. FA and LW equally contributed as shared first, YT and JK equally contributed as shared senior authors.

## Acknowledgements

FA is supported by the Berta-Ottenstein-Programme for Clinician Scientists, Faculty of Medicine, University of Freiburg. LW is supported by the Else Kröner-Fresenius-Stiftung (2016_Kolleg.03), Bad Homburg, Germany. ENH is supported by the Berta-Ottenstein-Programme for Advanced Clinician Scientists, Faculty of Medicine, University of Freiburg. YT is supported by the Else Kröner-Fresenius-Stiftung (2017_EKES.34), Bad Homburg, Germany and the Deutsche Forschungsgemeinschaft (SFB 1160 (P06/ B08)), Bonn, Germany.

Our deepest sympathy goes to all our patients and their relatives who were severely affected by COVID-19. We also want to express our sincere gratitude to our fellow health-care professionals who are providing outstanding patient care during this global health crisis, despite considerable personal risk.

